# A common model for the breathlessness experience

**DOI:** 10.1101/2020.09.29.20203943

**Authors:** Sarah L. Finnegan, Kyle T.S. Pattinson, Josefin Sundh, Magnus Sköld, Christer Janson, Anders Blomberg, Jacob Sandberg, Magnus Ekström

## Abstract

**Introduction:** Chronic breathlessness occurs across many different diseases, independently of severity. Yet, despite being strongly linked to adverse outcomes, chronic breathlessness is generally not considered a stand-alone treatment target. Here we move focus from identifying the “best” measurement tool and use data-driven techniques to identify and confirm the stability of underlying features (factors) driving breathlessness across different cardiorespiratory diseases. Such frameworks could provide an opportunity to address the underlying mechanisms of breathlessness and over-come issues with co-morbidities, particularly when medical therapies have been optimised.

**Methods:** Longitudinal study of questionnaire data on 182 participants with main diagnoses of asthma (21.4%), COPD (24.7%), heart failure (19.2%), idiopathic pulmonary fibrosis (18.7%), other interstitial lung disease (5.5%), and “other diagnoses” (8.8%) were entered into an exploratory factor analysis (EFA). Participants were stratified based on their EFA factor scores, allowing us to examine whether the breathlessness experience differed across disease diagnosis. We then examined model stability after six months and established through an iterative process the most compact, and therefore least burdensome assessment tool.

**Results:** From the 25 input measures, 16 measures were retained for model validation. The resulting model contained four factors to which we assigned the following descriptive labels: body burden, 2) affect/mood, 3) breathing burden and 4) anger/frustration. Stratifying patients by their scores across the four factors revealed two groups corresponding to high and low burden. These were not found to be predictive of primary disease diagnosis and did remain stable after six months.

**Conclusions:** We have identified four stable and disease-independent factors that seem to underlie the experience of breathlessness. We suggest that interventions may target factors within this framework to answer the question of whether they are also driving the experience itself.

## Introduction

Chronic breathlessness – breathlessness persisting despite optimal treatment, is a central symptom in many conditions, especially in respiratory and cardiac diseases, but also in cancer, neurological diseases and for survivors of COVID-19 [1, 2]. The impact of chronic breathlessness extends pervasively into people’s lives and leads to substantial personal, social and economic costs for the millions of sufferers world-wide [3]. Breathlessness is strongly linked to poorer clinical outcomes, including a higher number of hospital admissions and adverse events, poorer quality of life and increased rates of anxiety and depression [1, 4, 5]. While cardiorespiratory physiological mechanisms undoubtably often play a key role in breathlessness, they fail to explain breathlessness in many situations, such as in panic disorder, or when two individuals with objectively similar disease severities report very different experiences of breathlessness [5, 6]. Unlike fields such as pain, in which “chronic pain” is a stand-alone treatment target irrespective of the physical cause, no such equivalent exists for chronic breathlessness [1, 7]. These discrepancies, alongside the multifaceted and subjective nature of chronic breathlessness, make its assessment and treatment challenging. However, given that reported breathlessness is often a better predictor of short- and long-term survival than physiological measurements [4, 7-10], a mechanistic understanding of the experience would directly impact patient outcomes.

A multitude of assessment tools exist to quantify breathlessness. However, the focus is often on identifying the “best” measurement tool rather than underlying features (or factors) driving the experience of breathlessness. These factors may not only form the basis of a common descriptive framework for breathlessness but could also become key therapeutic targets. Similar approaches have already been used to identify baseline factors, which together predict treatment response in depression [11, 12] and pain [13, 14]. Our previous work has drawn upon machine learning techniques in order to search for common features both across assessment tools and across the individuals who complete them [15-17]. The separable factors revealed by this work centre around mood/affect measures [15-17] and symptom burden measures [15, 16], with further important factors including anticipated and physical capability measures [16]. Other independent studies using machine learning techniques to identify symptom-based phenotypes in asthma [18] and COPD [19, 20] also identified clusters of patients for whom breathlessness was linked with underlying mood/affect.

While our previous work has focused on identifying factors underling breathlessness within a single disease [16] or between a patient and a control group [15, 17], we have yet to determine whether a common model for the breathlessness experience can be identified across different cardiorespiratory diseases. Also, the stability of such models over time is unknown. Using a well characterised longitudinal dataset [21] of 182 patients with asthma, COPD, heart failure, idiopathic pulmonary fibrosis, other interstitial lung disease and “other diagnoses” including depression, cancer, diabetes and renal failure we addressed the following aims:

- *Establish whether a shared description of breathlessness is identifiable across disease diagnoses*
- *Determine if weightings on any identified shared factors would predict primary disease diagnosis*
- *Examine the stability of factors across time*
- *Establish the simplest informative model*

## Methods and Materials

This was an analysis of data from a longitudinal study of patients suffering from cardiorespiratory disease and breathlessness in everyday life. This body of work uses the dataset of which parts were used in published validation of the Swedish Multidimensional Dyspnea Profile (MDP) [21], the Dyspnoea-12 [22], and the instruments’ clinical feasibility and minimal clinically important differences [23]. The present analyses are novel and not previously reported.

### Participants

182 participants (97 female, median age 72 years [range 19-91 years], asthma (21.4%), COPD (24.7%), heart failure (19.2%), idiopathic pulmonary fibrosis (18.7%), other interstitial lung disease (5.5%), and “other diagnoses” including depression, cancer, diabetes and renal failure (8.8%), (Table 1)) were recruited from five outpatient clinics [21]. Inclusion criteria were: age 18 years or older, documented physician-diagnosed chronic cardiorespiratory disease, self-reported breathlessness during daily life defined as an answer “yes” to the question “Did you experience any breathlessness during the last 2 weeks?” and ability to give written informed consent to participate in the study. Exclusion criteria were: inability to write or understand Swedish adequately to participate, cognitive or other inability to participate in the study, or estimated survival of less than 3 months. Of the 182 participants who completed the baseline visit, 144 (79%) provided follow-up data at six months (79 female, median age 72 years [range 20-92 years]).

**Table 1.**
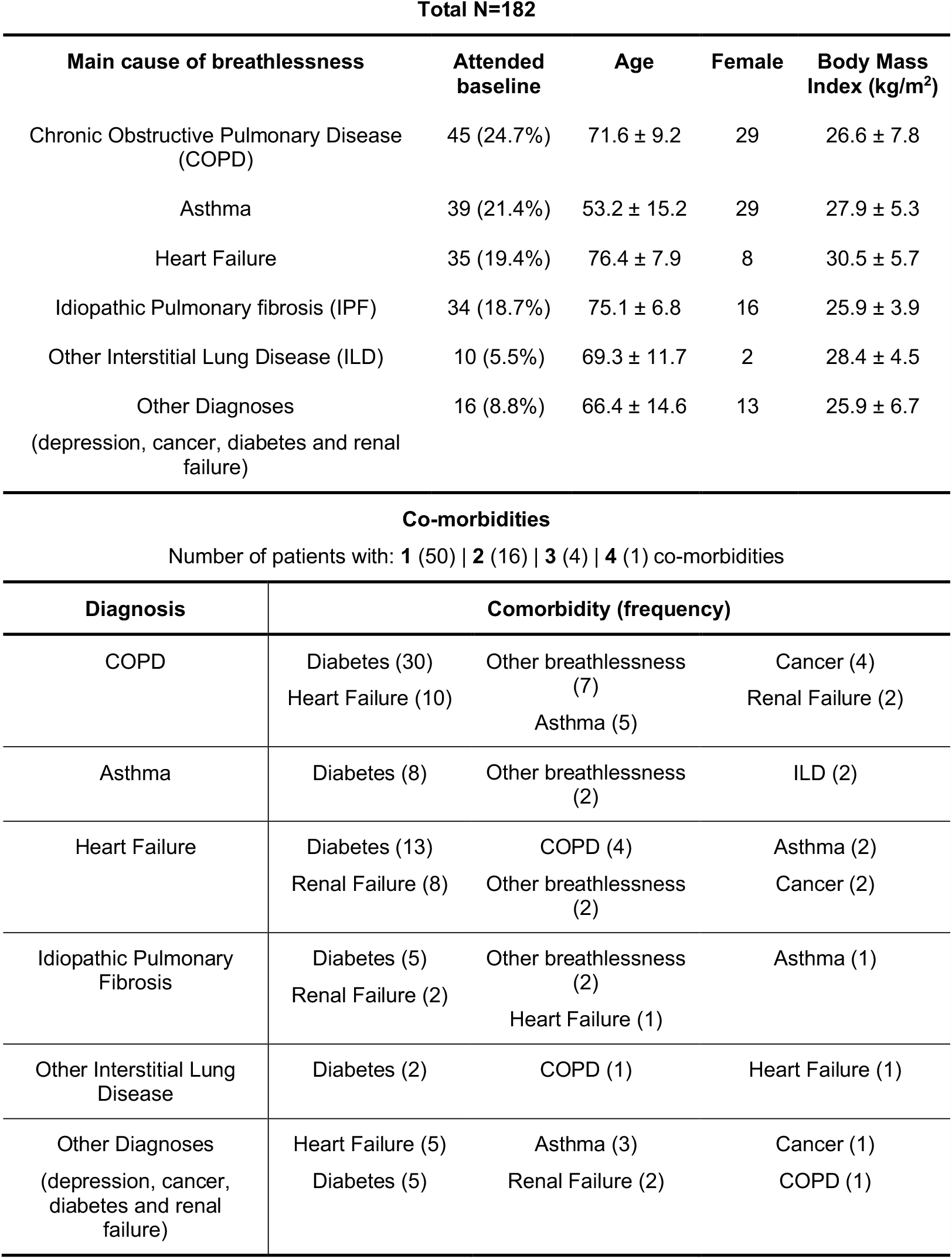
Demographic information expressed per group as mean ± SD or as frequency (percentage) and comorbidities expressed as frequency per main cause of breathlessness.

| Main cause of breathlessness | Attended baseline | Age | Female | Body Mass Index (kg/m<sup>2</sup>) |
| --- | --- | --- | --- | --- |
| Chronic Obstructive Pulmonary Disease (COPD) | 45 (24.7%) | 71.6 ± 9.2 | 29 | 26.6 ± 7.8 |
| Asthma | 39 (21.4%) | 53.2 ± 15.2 | 29 | 27.9 ± 5.3 |
| Heart Failure | 35 (19.4%) | 76.4 ± 7.9 | 8 | 30.5 ± 5.7 |
| Idiopathic Pulmonary fibrosis (IPF) | 34 (18.7%) | 75.1 ± 6.8 | 16 | 25.9 ± 3.9 |
| Other Interstitial Lung Disease (ILD) | 10 (5.5%) | 69.3 ± 11.7 | 2 | 28.4 ± 4.5 |
| Other Diagnoses<br>(depression, cancer, diabetes and renal failure) | 16 (8.8%) | 66.4 ± 14.6 | 13 | 25.9 ± 6.7 |

| Diagnosis | Comorbidity (frequency) |  |  |
| --- | --- | --- | --- |
| COPD | Diabetes (30)<br>Heart Failure (10) | Other breathlessness (7)<br>Asthma (5) | Cancer (4)<br>Renal Failure (2) |
| Asthma | Diabetes (8) | Other breathlessness (2) | ILD (2) |
| Heart Failure | Diabetes (13)<br>Renal Failure (8) | COPD (4)<br>Other breathlessness (2) | Asthma (2)<br>Cancer (2) |
| Idiopathic Pulmonary Fibrosis | Diabetes (5)<br>Renal Failure (2) | Other breathlessness (2)<br>Heart Failure (1) | Asthma (1) |
| Other Interstitial Lung Disease | Diabetes (2) | COPD (1) | Heart Failure (1) |
| Other Diagnoses<br>(depression, cancer, diabetes and renal failure) | Heart Failure (5)<br>Diabetes (5) | Asthma (3)<br>Renal Failure (2) | Cancer (1)<br>COPD (1) |

### Measures

Participants attended the clinic for a baseline visit, while repeat data were collected six months after the first visit date via a postal questionnaire. At baseline, demographic information including smoking status was collected. Participants completed the following self-report questionnaires, which were scored according to their respective manuals and recorded as their appropriate domain scores: COPD Assessment Test (CAT) [24]; Dyspnea-12 (D12) [3]; EuroQol Five Dimensions – Five Levels (EQ-5D-5L) [25]; Functional Assessment of Chronic Illness Therapy (FACIT)-Fatigue Scale; Hospital Anxiety and Depression Scale (HADS) [26]; Multidimensional Dyspnea Profile (MDP) [27]; modified Medical Research Council Breathlessness Scale (mMRC) [28]. Average severity of pain (0-10 NRS) and average severity of breathlessness were measured as “on average during the last two weeks” (Likert scale), along with current severity of breathlessness (0-10 numerical rating scale [NRS]). Baseline characteristics included measured height and weight.

Six months after their first visit participants were asked to complete and return a postal questionnaire pack. Questionnaires remained the same as at baseline and participants were asked to additionally rate their change in breathlessness since the first assessment on a 7-point ordinal scale (Global Impression of Change (GIC); where 1 = “very much better”, 4 = “no change” and 7 = “very much worse”) [23].

### Analysis

#### Factor identification

#### Aim to be addressed: establishing a shared description of breathlessness across disease diagnoses

Exploratory factor analysis (EFA) was used to identify and formalise any common structure underlying responses across clusters of questionnaire measures (Table 2). EFA is a model-free process, allowing researchers to examine a dataset without applying a preconceived structure to the result [29-31]. Via a three-step process, measures were grouped or discarded depending on how much they contributed to any one cluster. The resulting composite scores of each group were classed as a factor. Firstly, a parallel analysis with oblique rotation was employed to calculate the number of hidden (latent) factors within the dataset. Secondly, the number of questionnaire measures to retain was established. A maximum likelihood approach was applied, and measures that did not contribute (load at or above 0.4) onto a factor or demonstrated significant cross loading (i.e. loading onto more than one factor) were removed iteratively from the model. Finally, the model fit was tested by comparing the proposed model to a null model. Model selection criteria included variable loading at or above 0.4 with no cross loading or freestanding variables, a significant Tucker-Lewis Index (TL-index) close to 1, Root Mean Square Error of Approximation (RMSEA) < 0.06 and Standardised Root Square Mean Residual (SRMR) < 0.08. Models were fit using Lavaan version 0.6-1 in RStudio version 1.2.1.

**Table 2.** A list of measures included in the exploratory factor analysis model. BMI, Body Mass Index; D12, Dyspnea-12; MDP, Multidimensional Dyspnea Profile; SQ, Sensory Dimension; A2, Emotional Response Domain; HADS, Hospital Anxiety and Depression Scale; FACIT, Functional Assessment of Chronic Illness Therapy - Fatigue Scale; mMRC, modified Medical Research Council Breathlessness Scale; CAT, COPD Assessment Test; EQ5D, EuroQol Five Dimensions – Five Levels

|  |  |
| --- | --- |
| 1. Sex | 14. mMRC |
| 2. Age | 15. EQ-5D-5L – Index |
| 3. BMI | 16. FACIT - Fatigue Scale |
| 4. Dyspnea-12 – Affect | 17. Average severity of pain |
| 5. Dyspnea-12 – Physical | 18. CAT |
| 6. MDP_A2_Anxiety | 19. MDP_A2_Depression |
| 7. MDP_A2_Afraid | 20. MDP_A1_Breathing Discomfort |
| 8. MDP_A2_Angry | 21. MDP_SQ1_Physical Effort |
| 9. MDP_A2_Frustrated | 22. MDP_SQ2_Hunger |
| 10. Breathlessness at rest | 23. MDP_SQ3_Tightness |
| 11. Breathlessness mean | 24. MDP_SQ4_Mental Effort |
| 12. HADS - Depression | 25. MDP_SQ5_Hypernoea |
| 13. HADS – Anxiety |  |

### Participant stratification

#### Aim to be addressed: whether weightings on any identified shared factors predict primary disease diagnosis

Following exploratory factor analysis, each participant received a score (similar to the first component of a principle component analysis across measures within a factor) corresponding to each latent factor. To examine whether natural groupings of participants existed, the participants were stratified based on their factor scores using hierarchical cluster modelling techniques [32, 33]. Hierarchical models were used to reorder participants based on their correlation strengths [32]. Using unsupervised machine learning techniques, participants were linked firstly into pairs and then larger groups while fulfilling a cost function which equates to keeping within-group difference as low as possible while keeping between-group difference as large as possible. In this instance the algorithm would group two participants together who scored very similarly across factors, while placing individuals who scored dissimilarly into a second group. As the algorithm works in a hierarchical manner, individuals are represented at the bottom level, and participant clusters are at the top. The most distinct branching point for the hierarchical tree was determined by Matlab’s *evalclusters* function (MATLAB R2018b, Mathworks, Natick, MA). To determine whether the hierarchical groupings corresponded to disease diagnosis, the percentage probability of each of the disease categories was calculated for each group proportional to group size and compared with chance as shown in Equation 1.

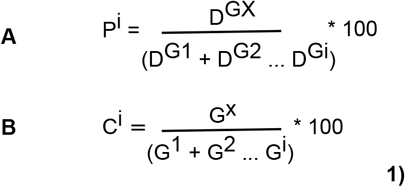

**Equation 1**. (***A*)** *For each group revealed by the hierarchical cluster model, the cause of breathlessness is calculated as a percentage of its total number of cases*. **(*B*)** *Chance is calculated proportionally to group size for each cause of breathlessness. Where P*^*i*^ *–percentage of cases of a cause of breathlessness, D*^*GX*^ *– number of a particular cause in a group, D*^*G1*^, *D*^*G2*^, *D*^*Gi*^ *– number of a particular cause across all groups* (*1,2*..*i*), *C*^*i*^ *– chance level calculated for each group identified by the hierarchical cluster model, G*^*X*^ *– number of individuals in group X, G*^*1*^, *G*^*2*^, *G*^*i*^ *– number of individuals across all groups*.

This method of establishing chance takes into account both potential differences in the sizes of groups arising from the hierarchical cluster model and differences in the numbers of participants with each diagnosis. Where A > B, individuals are considered to have an above chance probability of belonging to a particular group.

### Longitudinal model validation

#### Aim to be addressed: whether the factors remain stable across time

To determine whether the exploratory factor analysis model established at baseline was stable six months later we re-examined the factor structure using a confirmatory factor analysis on the follow-up data. Model fit criteria compared the proposed model to a null model. Model selection criteria were variable loading at or above 0.4 with no cross loading or freestanding variables, a significant Tucker-Lewis Index (TL-index) close to 1, Root Mean Square Error of Approximation (RMSEA) < 0.06 and Standardised Root Square Mean Residual (SRMR) < 0.08.

### Building a compact tool

#### Aim to be addressed: establishing the simplest informative model

Following the generation of the factor model we assessed whether low loading items could be removed from the model while maintaining a significant model fit. Model selection criteria were variable loading at or above 0.4 with no cross loading or freestanding variables, a significant Tucker-Lewis Index (TL-index) close to 1, Root Mean Square Error of Approximation (RMSEA) < 0.06 and Standardised Root Square Mean Residual (SRMR) < 0.08. The process was carried out iteratively and after each item’s removal the model fitting procedure was rerun and assessed for significance using the above model selection criteria. Items were removed until the model no longer significantly fit.

## Results

### Establishing a shared description of breathlessness across disease diagnoses

Of the 25 measures entered into the exploratory factor analysis (Table 2), 16 fulfilled the model fit criteria and were retained for model validation. The measures retained were: the EQ-5D-5L index, average severity of pain (0-10 NRS), (FACIT)-Fatigue Scale, CAT total score, HADS anxiety, mMRC, D12 physical, current and average severity of breathlessness (0-10 NRSs), MDP A1 (breathing discomfort), MDP A2 (frustration, anger, fear, anxiety and depression), MDP SQ4 intensity (mental breathing effort). The composite scores of the 16 retained measures formed four factors, which were validated according to the model fit criteria (TLI = 0.97, RMSEA = 0.055, SRMR = 0.03), shown in Figure 1:

**Figure 1.**
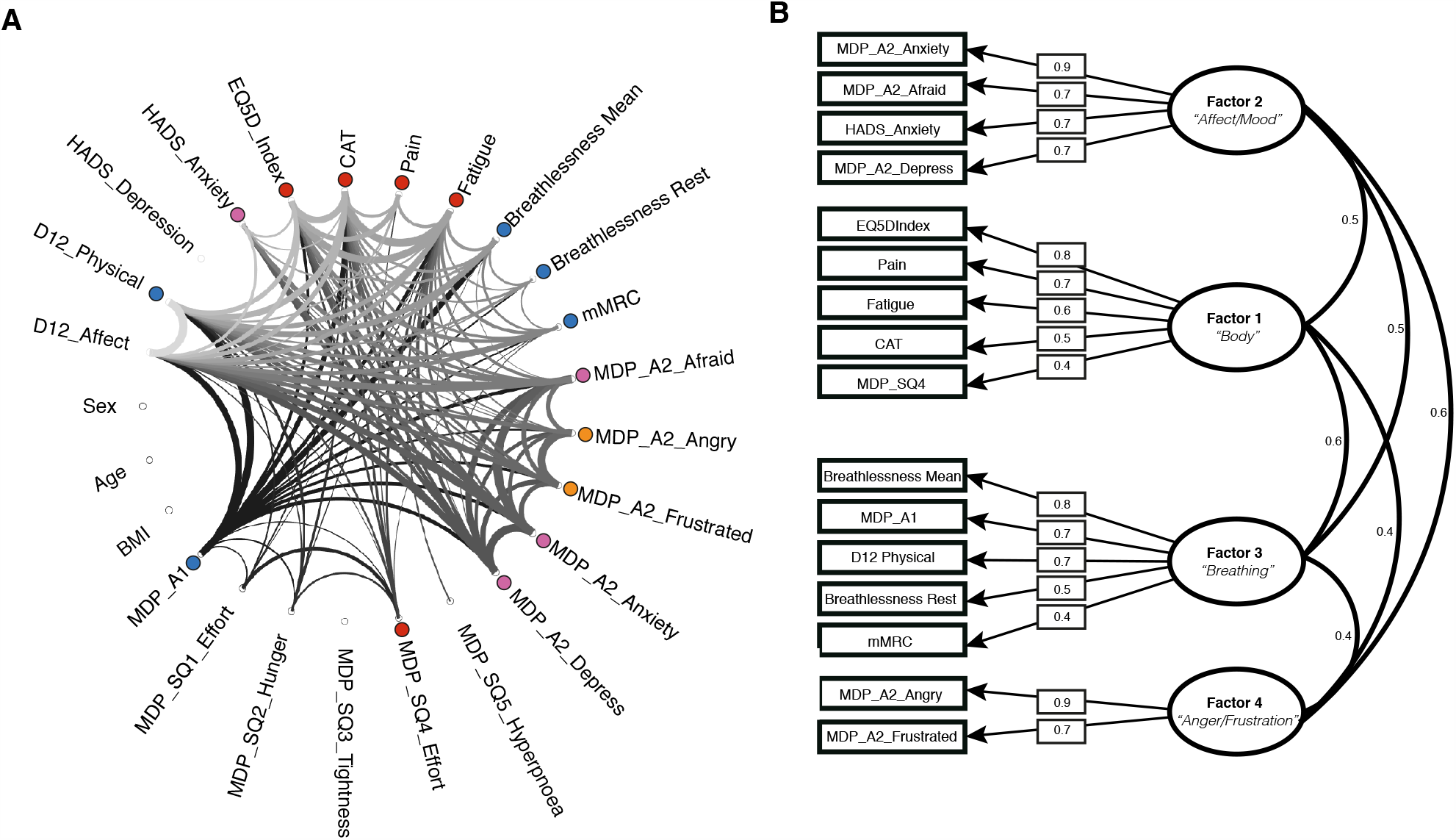
Connectogram. **(A)**–Measures are plotted as a wheel with connections between each measure input to the model shown as a thread. The thickness of the thread provides a visual representation of the strength of the relationship between any two measures (R>=0.35). Coloured dots highlight the common factors underlying groups of measures. Pink dots correspond to Factor 2 (affect/mood), red to Factor 1 (body burden), blue to Factor 3 (breathing burden) and orange to Factor 4 (anger/frustration). **Factor diagram (B)** The retained measures are shown on the left hand side of the diagram while the contribution of each measure to the resulting factor is shown as a loading weight (rectangular boxes) Factors are shown as ovals with descriptive labels underneath. The covariance across factors is illustrated by the curved lines.

- **Factor 1** – **Body Burden:** comprised EQ-5D-5L-index, pain and fatigue measures, CAT (a disease impact measure), and the SQ4 domain of the MDP (mental breathing effort).
- **Factor 2** – **Affect/Mood:** was composed of the anxiety, fear and depression A2 domains of the MDP along with the anxiety subscale of HADS.
- **Factor 3** – **Breathing Burden:** was made up of breathlessness at rest and on average, the mMRC, A1 domain of the MDP and the physical domain of the D12.
- **Factor 4** – **Anger/Frustration:** consisted of two of the A2 domains of the MDP – anger and frustration.

Factors 1 and 3 and Factors 2 and 4 showed the strongest covariance (curved lines Figure 1).

### Do weightings on any identified shared factors predict disease diagnosis?

Participants were stratified based on their four composite factor scores from the EFA model fit. A two-group solution was confirmed by MATLAB’s *evalcluster* algorithm as the most distinct and largely seems to correspond to high and low load across the four factors (Figure 2).

**Figure 2.**
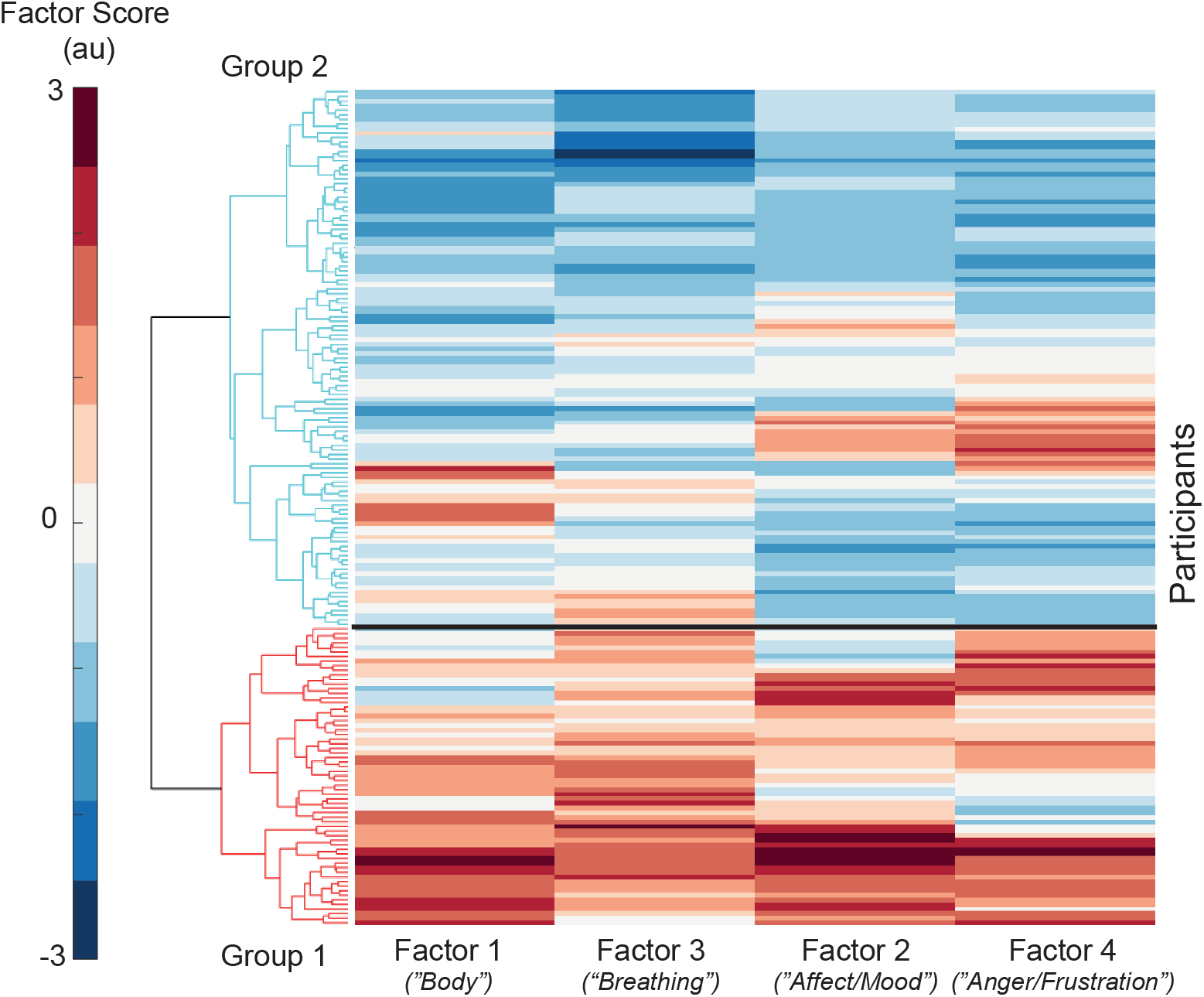
Clustergram: A matrix of each participants score across the 4 factors. Factor score is measures in arbitrary units (au). Each of the 4 factors is displayed along the bottom, with participants forming the y-axis. Factors 1 and 3 correspond to body and breathing burden, while Factors 2 and 4 correspond to affect/mood and anger/frustration. A dendrogram is displayed along the left side and highlights the division of participants into the two groups, where Group 1 – high symptom burden and Group 2 – lower symptom burden.

The two groups were not found to correspond to primary disease diagnosis, of which there were six categories asthma, COPD, heart failure, idiopathic pulmonary fibrosis (IPF), other interstitial lung disease and “other” (including depression, cancer, diabetes and renal failure). However, participants with IPF, other interstitial lung disease or heart failure were more likely when compared to chance (65%, 60% and 61% respectively) to be classified into Group 2 the “low load group”. While participants with COPD, asthma or a diagnosis of “other” were more likely to be classified into Group 1 the “high load group” when compared to chance (43%, 50% and 47% respectively).

#### Assessing the stability of factors over time

After six months the factor model was found to have remained stable according to the confirmatory factor analysis model fit criteria (TLI = 0.92, RMSEA = 0.086 (marginal fit), SRMR = 0.06).

#### Establishing the simplest informative model

The baseline model was then subjected to an iterative process in which the lowest loading variables were removed. After each cycle of variable removal, the model was retested. Figure 3 illustrates the final “compact” model. The MDP SQ4 (mental breathing effort) was removed along with mMRC, breathlessness at rest and CAT. The final model was found to be significant according to the model fit criteria (TLI = 0.96, RMSEA = 0.06, SRMR = 0.03), and was found to remain stable after six months (TLI = 0.93, RMSEA = 0.084, SRMR = 0.05), although RMSEA was considered to be marginal.

**Figure 3.**
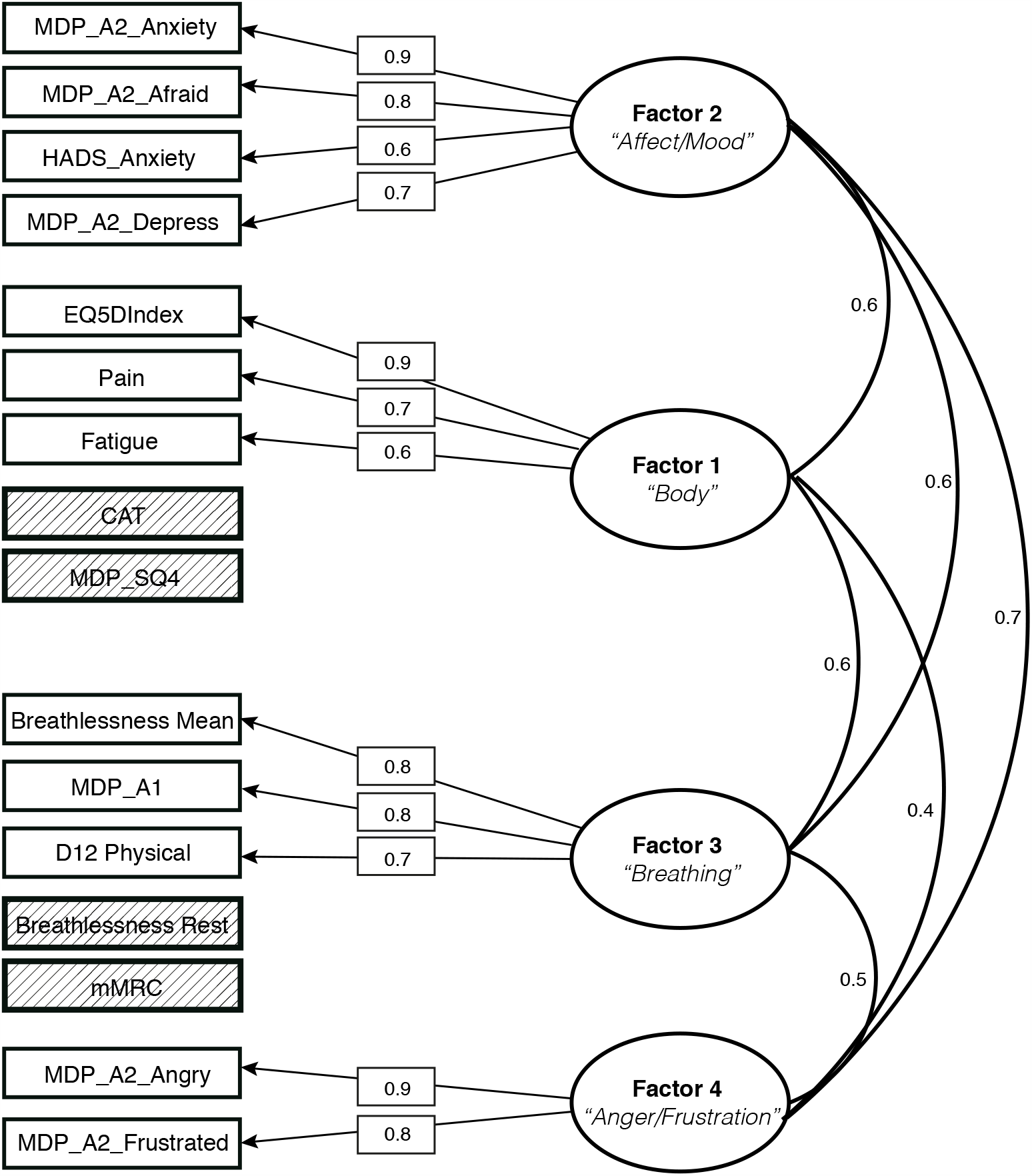
Factor diagram – a compact factor model in which low loading measures were sequentially removed. The retained measures are shown on the left-hand side of the diagram with the contribution of each measure to the resulting factor is shown as a loading weight (rectangular boxes). Removed measures are shown in cross-hatched boxes next to the factor to which they previously belonged. Factors are shown as ovals with descriptive labels underneath. The covariance across factors is illustrated by the curved lines.

## Discussion

### Key Findings

Using unsupervised machine learning techniques we identified four key factors underlying the experience of patients with chronic breathlessness. We assigned the key factors the following descriptive labels: body burden, affect/mood, breathing burden and anger/frustration. Together these factors provide a common description of breathlessness across asthma, COPD, heart failure, idiopathic pulmonary fibrosis and other interstitial lung disease. These factors were found to be stable across time but were not predictive above chance of the primary disease category. Instead participants fell into either high or low scorers across the four factors. Taken together, these findings go beyond our previous work by showing that there are common factors of breathlessness which are transferable across diseases and remain stable across time.

### Applying machine learning techniques to investigate breathlessness

In this study, we used unsupervised machine learning techniques, a benefit of which is that hypotheses and relationships can be explored without restricting or biasing the analytic process with investigator beliefs [29, 33]. This exploratory approach does not however, guarantee a statistically or clinically significant finding. Measures may have too little, or alternatively too much in common with all other measures to form separable factors. An example is the D12 affective score, which was removed from this model as it contributed strongly to both Factor 2 (mood/affect) and Factor 3 (breathing burden). These considerations lend confidence to our findings but reinforce the caveats of these techniques - factor analysis builds models based on shared variance and requires linear relationships between variables. Excluded variables not fulfilling those criteria may still be important descriptors of breathlessness. To address this, independent but relevant measures could be included at the point of participant stratification or as an independent validation of group differences.

### Latent factors of breathlessness

Parallels can clearly be drawn between the four factors identified in this study and our previous work despite different assessment tools being utilised. In an investigation of breathlessness in COPD we identified the most separable factors to be what a person felt they *could* or *could not* do, how their symptoms *impacted* their lives and their *general mood* [16]. Two of these factors, corresponding to mood and symptom burden, were identified in a second investigation conducted in individuals with asthma [15]. In this current work, mood/affect and symptom burden were again important factors, but here we were able to separate symptom burden into two factors; one focused on body burden (Factor 1) and a second factor relating to breathing burden (Factor 3). Interestingly, Factor 4, which contained anger/frustration measures did not collapse into Factor 2 (mood/affect), despite strong covariance. Both our previous and current work show measures contributing to factors corresponding to mood and body burden as relevant and distinct, while their strong covariance shows they are not completely independent. This illustrates the value of mechanistic research into this bi-directional relationship, which may become overlooked when investigated using other methods [6, 16, 34].

### Stratifying patients across diseases

Taking scores across the four-factor model we were able to split the participant population into two groups. One corresponding to higher scores across the four factors, and one lower scoring group. Again, this is consistent to our previous work which also found a two group structure corresponding to high and low scores across four factors [15, 16]. The current work extends our previous findings, as group identity was found to be independent of primary disease diagnosis, highlighting that a common psychological reference frame for breathlessness burden could provide an opportunity to address the underlying mechanisms of breathlessness, over-come issues with co-morbidities and drive treatment forwards in a more effective and personalised manner, particularly when medical therapies have been optimised. Stratification and cluster-based techniques have been used to good effect in other more clinically minded studies. Haldar et al and Leung et al were able to identify several different asthma phenotypes using similar methods [18, 35], but their works were restricted to mainly cardiorespiratory measures such as dosage of inhaled corticosteroids, neutrophil count, chemokine levels and atopy markers in a single disease.

### A repeatable and compact measure of breathlessness

A key requirement of any model is that it is stable across time. With this in mind we repeated our assessment of the factor structure on data acquired after six-months using confirmatory factor analysis and found that the factor structure remained stable. Having ascertained that the model was stable, we then sought to determine whether we could remove measures to create a more compact, less burdensome assessment, while maintaining a significant model fit. The iterative process of variable removal revealed only SQ4 (mental breathing effort) and CAT could be removed from Factor 1 (body burden), while mMRC and breathlessness at rest could be removed from Factor 3 (breathing burden). The final compact model structure (Figure 3) remained significant after testing on the six-month dataset. These results provide an endorsement that the model is capturing stable characteristics and therefore could be used for interventional investigations.

### Further considerations and limitations

In this work we have identified stable factors common to different disease populations that we hypothesise capture important self-report aspects of breathlessness. However, before firm conclusions as to the utility of this model can be drawn, we must address several questions. Firstly, do different weightings across the factors link with or predict relevant outcome measures? Are there different mechanisms underlying group identity? and finally are these groups a basis for personalised treatment pathways? To answer these questions we would need more detailed outcome measures and in depth physiological characterisation. If factor scores were found to link with clinical outcomes a randomised interventional study could target elements of the four factors and examine change scores across the outcome measures.

Future models should also consider building in the multiple co-morbidities common to patients with chronic breathlessness. In this work, we were restricted by sample size, and so individuals were labelled according to only their primary diagnosis, thus restricting the investigation of co-morbidities influence on symptom burden. However, with a larger sample size it may be possible to examine whether particular co-morbidities affect group identity or factor weightings. Additionally, physiology may be an important contributor to any description of breathlessness or a relevant outcome measure, but of the measures collected, none were suitable for use across all the clinical groups. Those that were collected would have likely biased the model towards exclusively detecting disease, for example cardiac left ventricle ejection fraction would be more relevant for heart failure than asthma. The difficulty of incorporating physiology into such models highlights the potential for compact patient-reported tools in examining the drivers of breathlessness across disease diagnoses. With low burden assessments we might be able to learn more about the mechanisms of breathlessness without being confounded by the associated physiology, which does not adequately describe or predict suffering [3, 8].

## Conclusions

We have shown using machine learning techniques that a common structure consisting of four factors, which we have labelled as body, affect/mood, breathing and anger/frustration, underlies patient reports of breathlessness that are independent of primary disease diagnosis. This structure, which remains stable over time could be used in interventional studies focused on targeted domains of breathlessness.

## Data Availability

The data used for this analysis are part of the longitudinal dataset reported as: Ekstrom, M., et al., Validation of the Swedish Multidimensional Dyspnea Profile (MDP) in outpatients with cardiorespiratory disease. BMJ Open Respiratory Research, 2019. 6(1): p. e000381

## Acknowledgements

The authors extend their warm thanks to the staff conducting the study, to Hans Bornefalk and Anna Hermansson Bornefalk who made important contributions regarding the statistical aspects of the project and database management, and to all patients who participated to make this research possible.

